# Remote Assessment of Mental Health and Physical Activity in Older Black Americans during the COVID-19 Pandemic

**DOI:** 10.1101/2025.05.23.25328188

**Authors:** Ching-Hsuan Shirley Huang, Andrew Shutes-David, Sarah Payne, Katie Wilson, Karl Brown, Kevin M. Vintch, Edmund Seto, Debby W. Tsuang

**Affiliations:** Department of Environmental and Occupational Health Sciences, University of Washington, Seattle, WA, United States; Geriatric Research, Education, and Clinical Center, VA Puget Sound Health Care System, Seattle, WA, United States; Mental Illness Research, Education, and Clinical Center, VA Puget Sound Health Care System, Seattle, WA, United States; Department of Biostatistics, University of Washington, Seattle, WA, United States; Department of Psychiatry and Behavioral Sciences, University of Washington, Seattle, WA, United States

**Keywords:** COVID-19, remote assessment, mental health, Black Americans, aging

## Abstract

Older Black Americans (BAs) face disparities in diagnosing and treating cognitive and mental health conditions. The shift to remote care during the COVID-19 pandemic may have exacerbated these inequities through digital barriers. This pilot study evaluated the feasibility of remote mental-health assessments and accelerometry among older BAs (n=6, age 65+, with subjective memory complaints but no dementia diagnosis). Participants completed three 2-week assessment modalities: pen and paper, telephone, and online/videoconferencing, and wore wrist accelerometers to measure physical activity. All modalities were well-tolerated, with participants expressing the strongest preference for the personal contact of telephone and online/videoconferencing and enthusiasm for accelerometry. The UCLA-Loneliness score demonstrated significant positive correlations with GAD-7 and significant negative correlations with LSNS-R and SF-20 mental-health domain. Although significant correlations between physical activity levels and mental-health assessment scores were not observed, trends in correlation coefficients suggests that mean daily, daytime, and nighttime hourly activity counts were negatively correlated with GAD-7 and UCLA-L and positively with LSNS-R scores. These findings suggest older BAs are amenable to accelerometry and remote assessments involving personal contact with providers. Observed trends suggest physical activity may be associated with reduced anxiety, loneliness, and social isolation. Larger studies are necessary to confirm these potential findings.

## 1. Introduction

Compared to older white Americans (WAs), older Black Americans (BAs) are more likely to develop dementia ^1^ and encounter racial disparities related to diagnosis and care ^2–4^, to have undiagnosed ^5^ or untreated depressive symptoms ^6^, to have more severe anxiety disorders ^7^, to rate their mental health as worse ^8^, and to experience “digital and technical inequities” that reduce their access to, familiarity with, or acceptance of technologies ^9–11^. During the COVID-19 pandemic, older patients reported feeling socially isolated from support systems (e.g., community centers and churches) and bereft of mental-health support ^12^, which put them at greater risk for depression, anxiety, cognitive impairment, sleep disturbances, suicide, and death ^12–17^. Many of these risks were likely exacerbated for older BAs with cognitive impairment or dementia ^18^ and compounded due to BAs’ concerns related to technology, including, perhaps, the kinds of remote health care technology and virtual appointments that became ubiquitous during the socially distanced years of the pandemic 19.

Although the widespread use of remote technologies to perform virtual clinical visits may have ebbed somewhat since 2020, virtual clinic visits are still significantly more common for patients with dementia than before the pandemic ^20^, and they continue to represent an important tool for both patients and providers. It is thus essential that we consider ways to bridge the “digital divide” ^21^ and to confront racial disparities in the diagnosis and care of mental health disorders, particularly for older BAs with cognitive impairment.

In this small pilot study, we captured three kinds of preliminary data to help illuminate the role that technology and remote health assessments might or might not play in the characterization of mental health for older BAs. First, we used a within-subject design to compare three different modalities for remotely assessing mental health—pen and paper, telephone, and online/videoconferencing. Second, we explored the use of accelerometry, a form of passive monitoring technology that may avoid barriers related to technical knowledge and experience. This second approach was based on our sense that accelerometry devices that monitor physical activity may be well tolerated by older BAs and may provide useful data related to cognitive impairment and mental health. And third, we performed a series of satisfaction interviews to help characterize BA perspectives concerning these varying assessment modalities and the use of accelerometry. Together, the preliminary findings in this study provide a snapshot of how remote methodologies could be implemented in older BAs, as well as a snapshot of the relationships between mental health and physical activity within this underrepresented population.

## 2. Methods

### 2.1 Study design and subject recruitment

In Spring/Summer 2021, BA adults who were age 65+ and scored 20 or higher on the Montreal Cognitive Assessment (MoCA) (n=26) were mailed a recruitment letter and then enrolled (n=7) as part of a three-phase protocol that was approved by the VA Puget Sound Health Care System institutional review board. In phase A, participants completed mailed assessment packets that included the Generalized Anxiety Disorder scale-7 (GAD-7), Short Form Health Survey (SF-20) composite score, UCLA Loneliness Scale (UCLA-L), and Lubben Social Network Scale (LSNS-R); in phase B, participants completed the same assessments over the telephone; and in phase C, participants completed the same assessments online and as part of videoconferencing appointments. Participants were randomly assigned to a phase order, and each assessment modality was performed twice during each 2-week phase (except the SF-20, which was only completed once during each phase).

Participants also wore an accelerometry device on their nondominant wrist during each phase. On the last day of each phase and at the end of their study participation, a survey was administered to participants and study informants to capture their perceptions of the assessments and accelerometry methods. Each phase was separated by a washout period of at least 2 weeks.

### 2.2 Mental health assessments

Four mental health outcomes were assessed in each phase (see **Supplementary Table 1**): anxiety, perception of physical health, loneliness, and social engagement. As noted above, these outcomes were evaluated using standardized questionnaires that were administered either weekly or one time during each study phase. The SF-20 questionnaire assessed six subdomains: physical functioning, role functioning, social functioning, mental health, health perception, and pain. Within these domains, higher scores indicate better outcomes, except for the pain domain, in which a higher score reflects a worse outcome. For the GAD-7 and UCLA-L scales, higher scores correspond to elevated levels of anxiety and loneliness, respectively, indicating poorer outcomes. Conversely, for the LSNS-R scale, lower scores indicate higher levels of loneliness.

### 2.3 Physical activity measures

Accelerometer-based physical activity was assessed using the Actigraph GT3X+ (Actigraph Corporation, Pensacola, FL). Participants wore the device for 14 consecutive days during each phase. A sampling rate of 60 Hz with 60-second epochs was used. In the analyses, activity counts, calculated as the vector magnitude of the three axes (VMCounts), were used as the proxy for physical activity. To ensure valid data, wear and non-wear time were identified using a validated algorithm (90 consecutive minutes of zero counts were marked as non-wear, allowing for up to 2 minutes of nonzero counts). Data were included only if wear time exceeded 70% of the day (≥ 1,008 minutes).

Accelerometry data were then aggregated into 1-minute intervals and further summed into hourly VMCounts. To explore diurnal activity patterns, VMCounts and MVPA were calculated separately for daily (12 AM–11:59 PM), daytime (7 AM–8:59 PM), and nighttime (9 PM–6:59 AM) periods.

Correlations between different VMCounts metrics and mental health outcomes were examined using Spearman’s rank correlation coefficients. For this analysis, data from Phase A and Phase B were combined to ensure sufficient data pairs, whereas Phase C was excluded due to missing accelerometry data for two participants. Prior to correlation analysis, VMCounts metrics and mental health assessment scores were averaged within each phase to generate phase-level values for each participant.

### 2.4 Perception surveys

The perception surveys used both multiple choice and open-ended questions and were administered online. The surveys queried the clarity of assessment/device instructions; the level of support provided by the research team; the perceived effectiveness, difficulty, and cultural sensitivity of the assessments/device; the amount of assistance needed from the study informant or research staff to complete the tasks; and the participants’ interest in using these methods in the future with their health care providers. Participants were also asked which methods they preferred. Survey responses were analyzed using descriptive statistics for multiple-choice questions, whereas qualitative responses were reviewed thematically to identify key participant perceptions.

## 3. Results

### 3.1 Subject characteristics

Of the 7 participants recruited into the study, 1 took part in the modality assessments but had no physical activity data collected and was therefore excluded from the analyses. The remaining 6 participants were between the ages of 69 and 78 with a mean age of 72.7 (standard deviation, SD=4.9); primarily male (n=5); and had a mean MoCA score of 23 (SD=1.96) at enrollment, suggesting mild cognitive impairment; none of the participants had a clinical diagnosis of dementia.

### 3.2 Mental health

Mean participant scores are depicted in **Table 1**. Participant UCLA-L scores demonstrated a significant positive correlation with the GAD-7 and a significant negative correlation with the LSNS-R and the mental health domain of the SF-20 (see **Figure 1**). This may suggest that increased loneliness is associated with higher anxiety levels, decreased social engagement, and poorer mental health. The LSNS-R and SF-20 social domain scores were correlated such that higher levels of pain were associated with decreased social engagement (see **Figure 1**). This finding may suggest that individuals experiencing pain will engage in fewer social activities, which could further exacerbate feelings of loneliness and negatively impact overall mental well-being.

**Table 1.** Summary statistics of the mental health assessment scores and daily VMCounts by phases.

| Variables | Mean (SD) |  |  |
| --- | --- | --- | --- |
|  | Phase A | Phase B | Phase C |
| GAD-7 (n=6) | 1.7 (2.5) | 2.4 (3.1) | 2.5 (3.0) |
| SF-20 (n=6) | 59.0 (13.7) | 60.5 (9.3) | 68.9 (10.8) |
| LSNS-R (n=6) | 31.2 (10.4) | 30.0 (12.3) | 26.1 (12.0) |
| UCLA-L (n=6) | 34.3 (16.2) | 33.0 (15.2) | 28.3 (23.9) |
| Daily VMCounts* | $1.43 \times 10^6$ ($4.4 \times 10^5$) | $1.58 \times 10^6$ ($6.6 \times 10^5$) | $1.45 \times 10^6$ ($3.4 \times 10^5$) |

\* n=6 for Phase A and B, and n=4 for Phase C (due to missing accelerometry data for two participants in that phase).

**Figure 1.**
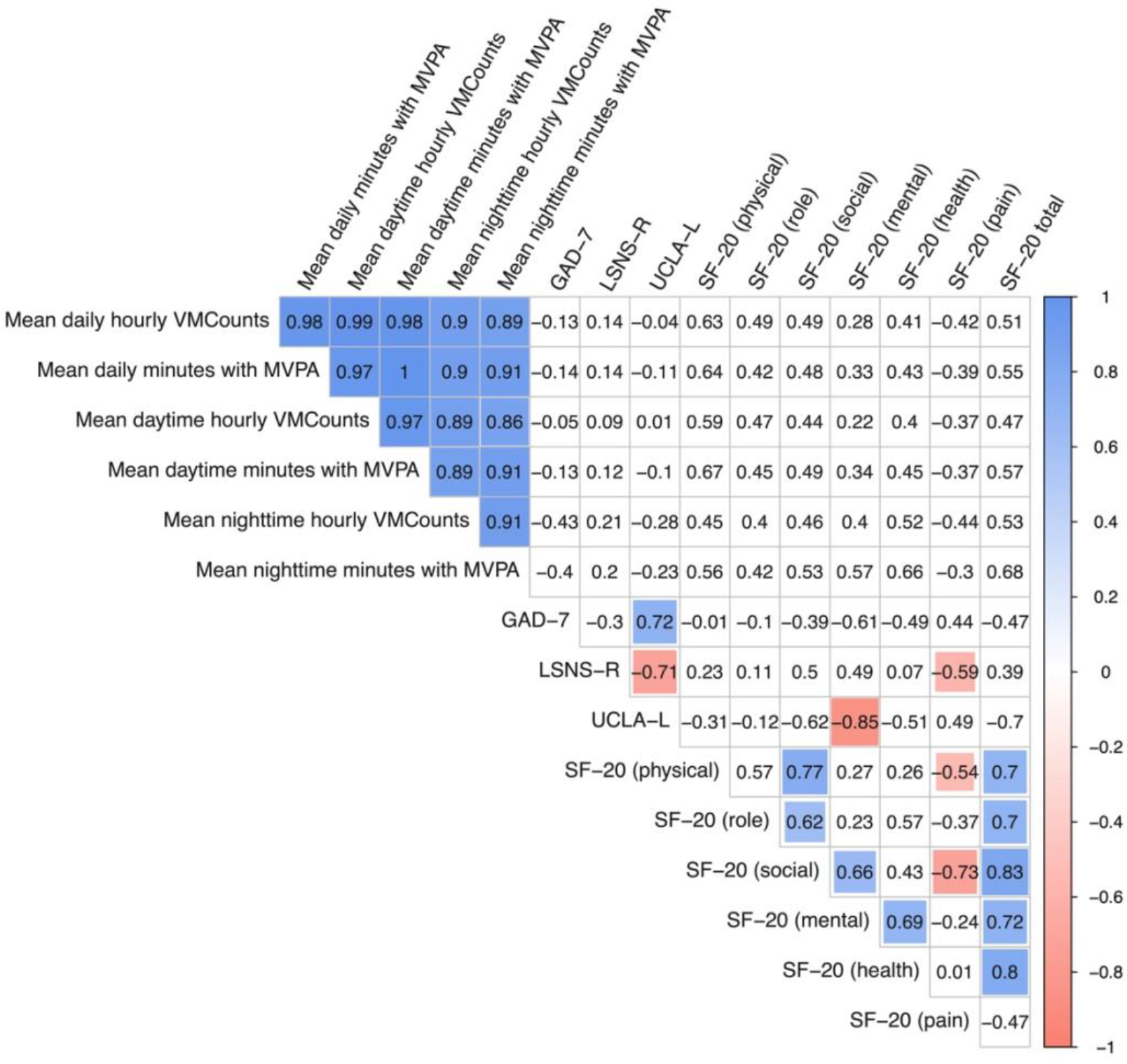
Spearman’s correlation coefficient Matrix: Relationship between mean VMCounts and mean minutes with MVPA (24-hr, daytime, nighttime) and GAD-7, LSNS-R, UCLA-L, and SF-20 subdomains. Phase A and Phase B data are combined for correlation coefficient calculation. Phase C data were excluded due to insufficient data points for pairwise correlation analysis. Colored cells indicate nominal p-values < 0.05.

### 3.3 Hourly physical activity level

Participants appeared to tolerate the accelerometry devices well, and no clear patterns were observed in wear or non-wear times during the study phases (see **Table 1** for mean daily VMCounts and **Supplementary Figure 1** for wear times, excluding days with less than 70% wear time).

We observed greater variability in hourly VMCounts during daytime hours (7 am – 8:59 pm) and less variability during the nighttime and early morning hours (9 pm – 6:59 am); this may indicate diverse activity patterns among participants during the day but periods of sleep or reduced activity at nighttime and early morning (see **Supplementary Figure 2**, which presents the variation in hourly VMCounts at each time of day [0 – 24 h] during a 14-day study phase).

The diurnal patterns of hourly VMCounts are consistent for each subject, with higher median hourly VMCounts recorded primarily during the daytime and early evening. Despite this consistency, some individuals exhibited activity extending beyond the typical daytime hours, as reflected by elevated hourly VMCounts until midnight. Generally, the pattern of hourly VMCounts was consistent across the different study phases for each participant.

We also found that the distribution of total hourly VMCounts appears to vary across different study phases and participants, suggesting individual variability (see **Supplementary Figure 3** for boxplots of the distribution of 24-hr, daytime, and nighttime total hourly VMCounts by participant across study phases). As expected, the median daytime hourly VMCounts were higher than the median nighttime hourly VMCounts for all participants, and in most instances, variability during the daytime hours tended to be higher than during the nighttime hours. However, we noted exceptions, such as with Participant #4 who, in Phase C, exhibited greater nighttime variability in hourly VMCount than daytime variability.

We also noted a similar pattern in regard to MVPA such that more daytime minutes were spent in MVPA and fewer nighttime minutes were spent in MVPA (see **Supplementary Figure 4**).

### 3.4 Relationship between physical activity level and mental health

As depicted in the correlation matrix (**Figure 1**), we observed trends between physical activity metrics and mental health outcomes that were consistent with expectations. More specifically, the mean daily, daytime, and nighttime hourly VMCounts were negatively correlated with GAD-7 and UCLA-L scores, suggesting that higher physical activity levels may be associated with reduced anxiety and loneliness, and positive correlations were observed between VMCounts metrics and LSNS-R scores, suggesting that higher physical activity levels may be correlated with increased social engagement. In addition, all of the SF-20 assessment domains except the pain domain (i.e., the physical, role, social, mental, and health domains) were positively correlated with physical activity levels, suggesting that higher physical activity may be associated with a better overall perception of physical health.

We also examined the relationship between physical activity and mental health by study phase (see **Supplementary Figure 5** and **6** for Phase A and Phase B, respectively).

Although some changes were observed in the direction and strength of correlations when comparing the phase-specific findings to the combined dataset findings (i.e., as depicted in **Figure 1**), no consistent patterns emerged to suggest phase-dependent effects. Some relationships that appeared stronger in the combined dataset became weaker when analyzed separately by phase, whereas others showed changes in direction. This variability suggests that these differences may be due to sample size limitations or random variation rather than meaningful phase-dependent effects.

### 3.5 Perception survey results

After completing the study, among the 6 participants, no one indicated a preference for the pen-and-paper assessments, 75% indicated a preference for telephone assessments, and 25% indicated a preference for the online and videoconferencing assessments. Participants who favored the telephone assessments explained that it was easier to remember to complete the forms during a scheduled phone call than during the other phases. Participants who preferred the online assessments described preferring face-to-face communication and disliking the telephone assessments because they took too long to complete; this was also the primary complaint about the pen-and-paper assessments.

In addition, the participants who wore the accelerometry device tolerated it well, with some expressing surprise at their own comfort with the device. Three participants specifically noted that wearing the device served as a reminder to increase their physical activity and make health-conscious choices.

## 4. Discussion

Numerous studies have investigated race-related disparities in the assessment and treatment of cognition and mental health, and many studies have sought to characterize minority views and usage patterns in regard to telehealth, online health instruments, and other health-related technologies. However, to our knowledge, no studies have specifically considered different remote modalities for administering mental health assessments in BAs—indeed, in a systematic review, McCall et al. ^22^ specifically note the lack of studies investigating different modalities for evaluating anxiety and depression in BAs.

In this small study, we found that BA participants preferred the personal contact of the telephone and online/videoconferencing mental health assessments over the pen-and-paper mental health assessment. They particularly highlighted the ways in which certain assessments were easier to remember to complete (i.e., the telephone assessments), provided more opportunities to connect with the research team (i.e., videoconferencing), or took less time to complete (i.e., online). This parallels the findings of Gamaldo et al. ^23^, who compared pen-and-paper cognitive assessments to computer-based cognitive assessments (i.e., the CogState and Joggle) in older BAs (age=55-86) and found that older BAs preferred the computer tests. Hoge et al. ^24^ also compared assessment modalities in BAs, but their study was markedly different; that is, they compared remote and in-person cognitive assessments in younger BAs (age=46.2; SD=11.9) with systemic lupus erythematosus.

Our analysis of remote mental health assessments also showed that loneliness was associated with anxiety, decreased social engagement, and poorer mental health for older BAs with subjective cognitive impairment. This finding is consistent with the findings of investigators who compared GAD-7, UCLA-L, and LSNS scores from the COVID era e.g., ^25^ but in more expansive populations.

Another important part of this study is our use of accelerometry in older BAs. Other investigators have performed accelerometry studies in BAs e.g., ^26^ but not necessarily by focusing on older BAs or on BAs with mild cognitive impairment. To our knowledge, this is also the first study to elicit BA perspectives concerning the use of accelerometry. We found that the accelerometry device was well-tolerated, that none of the participants who wore the device expressed reservations about wearing it for multiple 14-day sessions, and that some of the participants expressed enthusiasm for the ways in which wearing the device encouraged them to be more conscious about their health and activity choices.

Although our accelerometry findings were not subjected to formal significance testing due to the small sample size, we observed consistent trends suggesting that higher levels of physical activity may be associated with lower levels of loneliness, anxiety, and social disengagement, as well as more favorable perceptions of physical and mental health. These findings should be interpreted as exploratory and hypothesis-generating rather than conclusive. Nonetheless, these findings offer valuable insights into the interplay between physical activity and mental well-being among older BAs during the COVID-19 pandemic and beyond.

Finally, although the pandemic is “behind” us, its lasting impact on the mental health of older BAs remains a concern, particularly given the historical neglect of appropriate mental health assessments and care for BAs. This study suggests that telephone and online/video-conferencing assessments of mental health may be appropriate for older BAs, highlights the potential importance of physical activity in helping to ameliorate mental health symptoms and encourage social connectivity, and underscores the need for future studies with larger sample sizes within older BA populations to tease out the specific relationships between physical activity and mental health.

## Data availability statement

The datasets presented in this study are not publicly available due to restrictions related to confidentiality.

## Acknowledgments

We thank Ciara DeGraff for her work with study participants, as well as the volunteers who made this research possible.

## Author Contributions

SCH: Formal analysis, Methodology, Software, Visualization, Writing – original draft, Writing review & editing. ASD: Conceptualization, Funding acquisition, Writing – original draft, Writing – review & editing. SP: Data curation, Formal analysis, Investigation, Project administration, Writing – review & editing. KW: Conceptualization, Formal analysis, Writing – review & editing. KB: Formal analysis, Validation, Writing – review & editing. KMV: Data curation, Formal analysis, Investigation, Project administration, Writing review & editing. ES: Conceptualization, Formal analysis, Funding acquisition, Methodology, Supervision, Writing – review & editing. DWT: Conceptualization, Funding acquisition, Methodology, Project Administration, Resources, Writing – review & editing.

## Supplementary Material

**Supplementary Table 1.** Summary of cognitive and mental health questionnaires.

| Questionnaire | Outcome | Scoring | Assessment frequency |
| --- | --- | --- | --- |
| Montreal Cognitive Assessment (MoCA) | Cognition | $\geq 26$: no cognitive impairment<br>19–25: mild<br>10–18: moderate<br>$\leq 9$: severe cognitive impairment | At baseline |
| Generalized Anxiety Disorder scale-7 (GAD-7) | Anxiety | 0–4: minimal anxiety<br>5–9: mild<br>10–14: moderate<br>15–21: severe anxiety | Weekly during each phase |
| Short Form Health Survey SF-20 composite score | Perception of physical health | Varies by domains<sup>a</sup> | Once during each phase |
| UCLA Loneliness Scale (UCLA-L) | Loneliness | 20–34: low loneliness<br>35–49: moderate<br>50–64: moderately high<br>65–80: high loneliness | Weekly during each phase |
| Lubben Social Network Scale (LSNS-R) | Social engagement | $\leq 12$ denotes at risk for social isolation | Weekly during each phase |

<sup>a</sup> Measured 6 subdomains, including physical functioning, role functioning, social functioning, mental health, health perceptions, and pain.

**Supplementary Figure 1.**
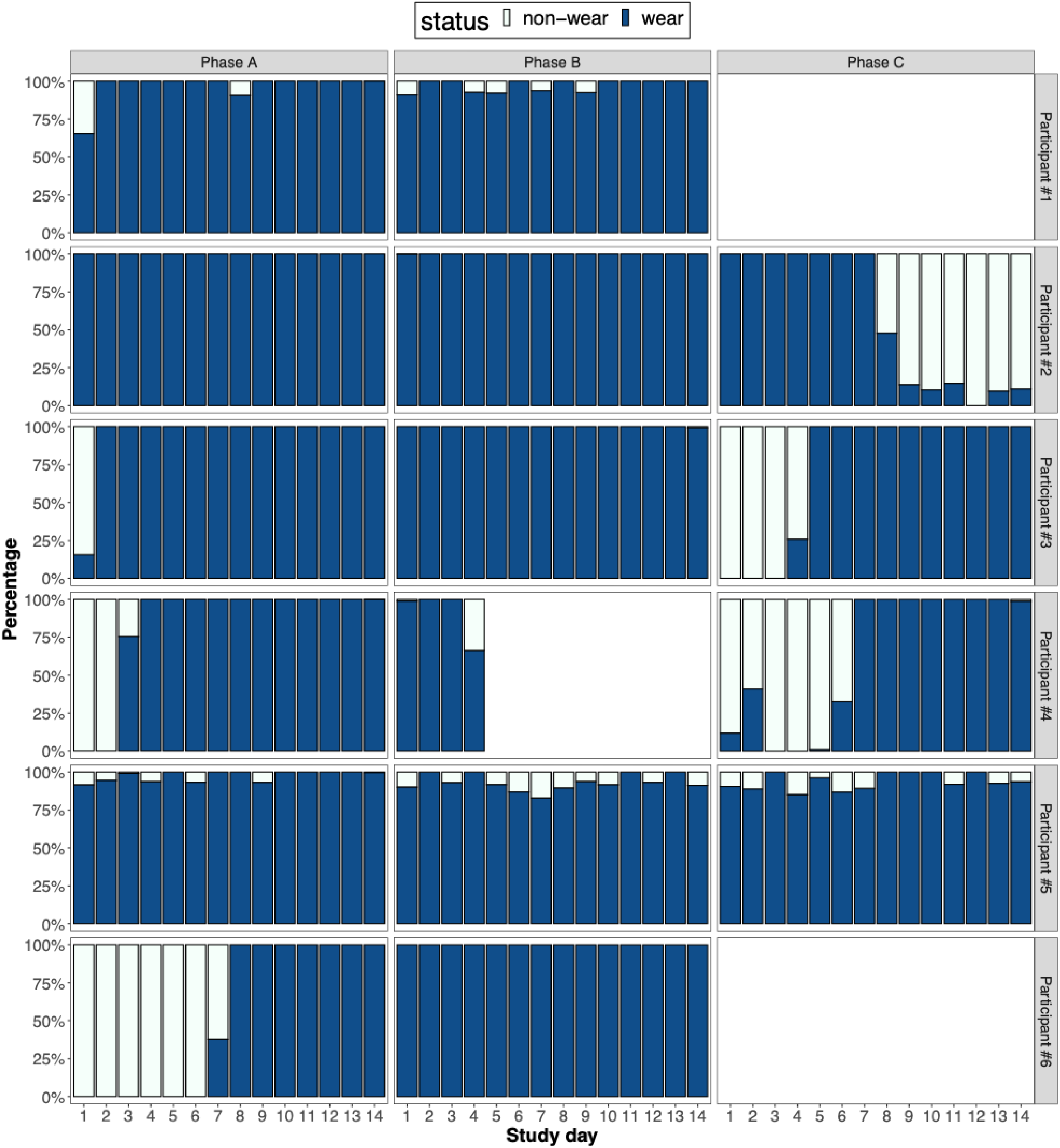
Accelerometry device wear vs. non-wear time by study phase.

**Supplementary Figure 2.**
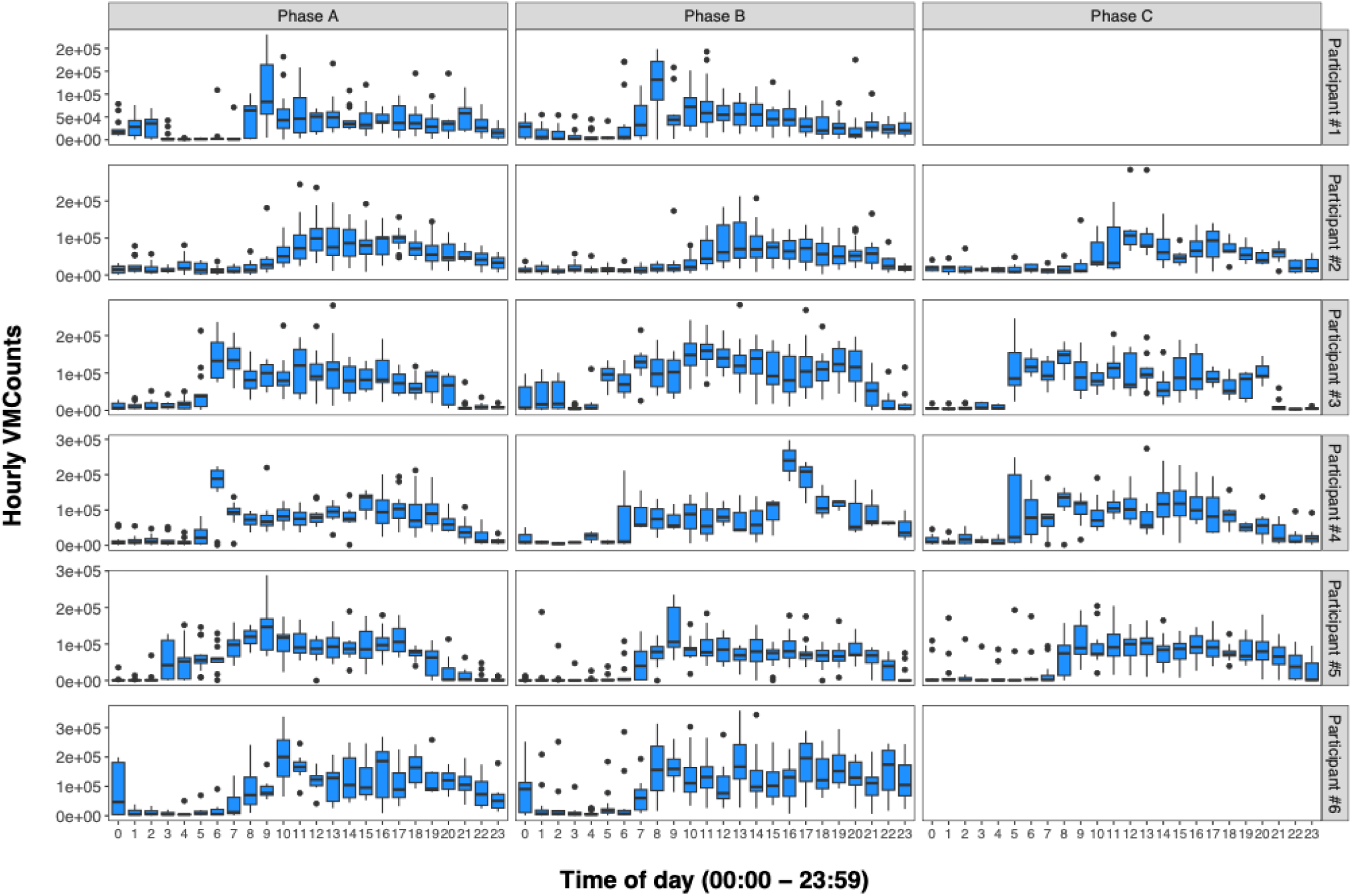
Boxplots of hourly VMCounts by time of day, stratified by participant and study phase. Each box shows the distribution of the hourly total VMCounts over specific times of day during the 14-day study period. Note the y axes are on a different scale for each participant.

**Supplementary Figure 3.**
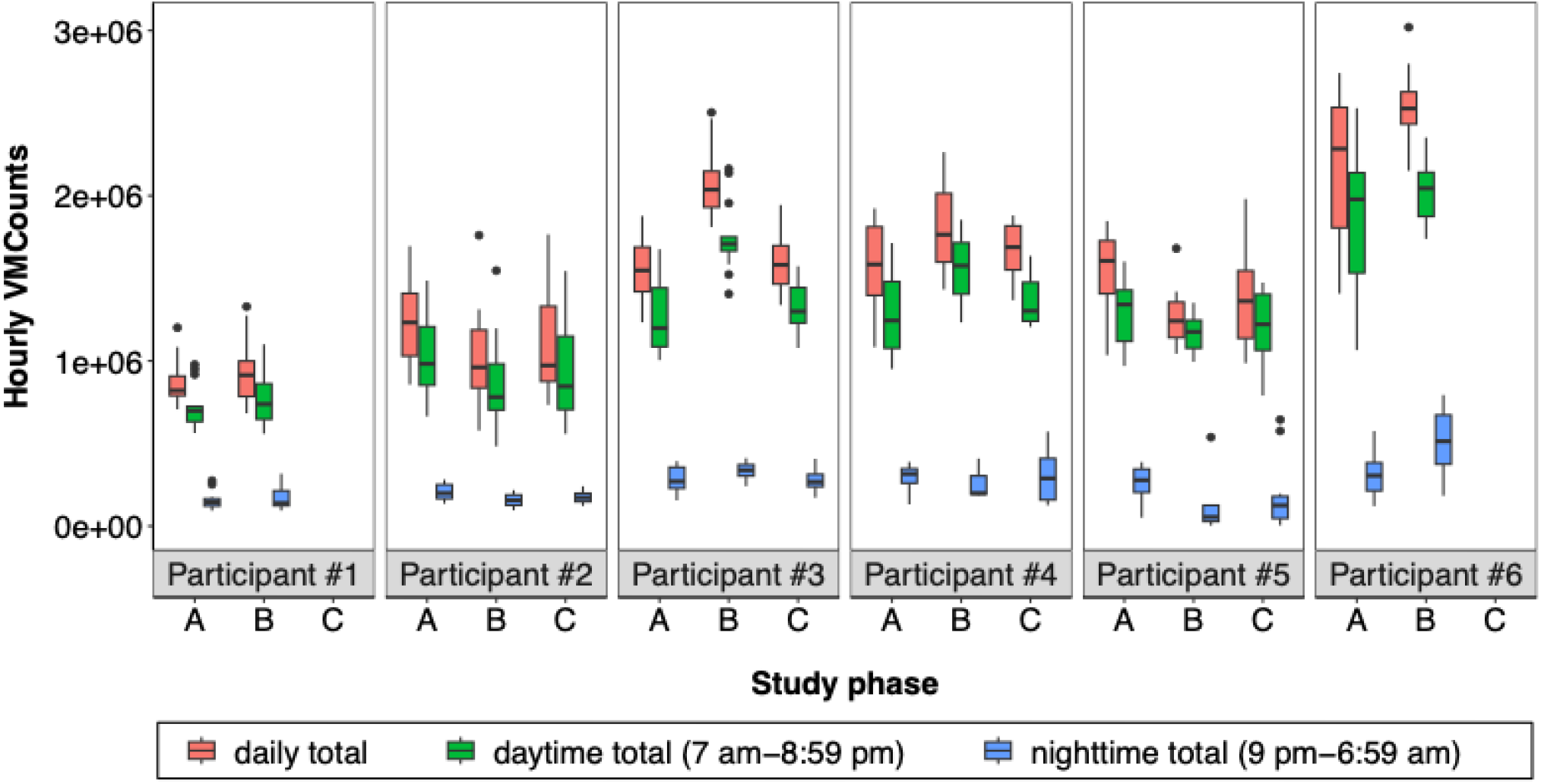
Boxplots of the distribution of 24-hr, daytime, and nighttime total hourly VMCounts by participant across study phases.

**Supplementary Figure 4.**
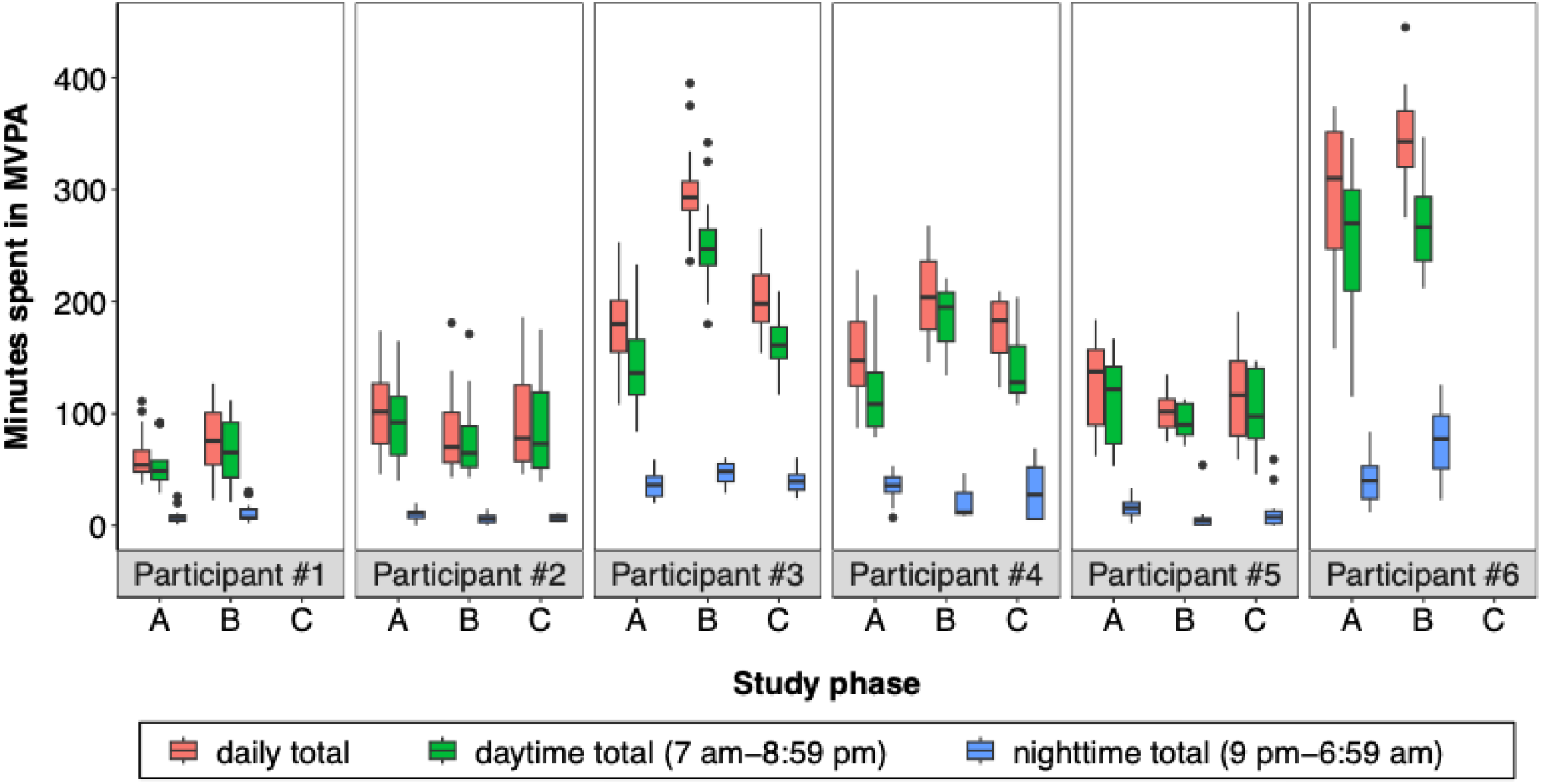
Boxplots of the distribution of 24-hr, daytime, and nighttime total minutes spent in MVPA by participant across study phases.

**Supplementary Figure 5.**
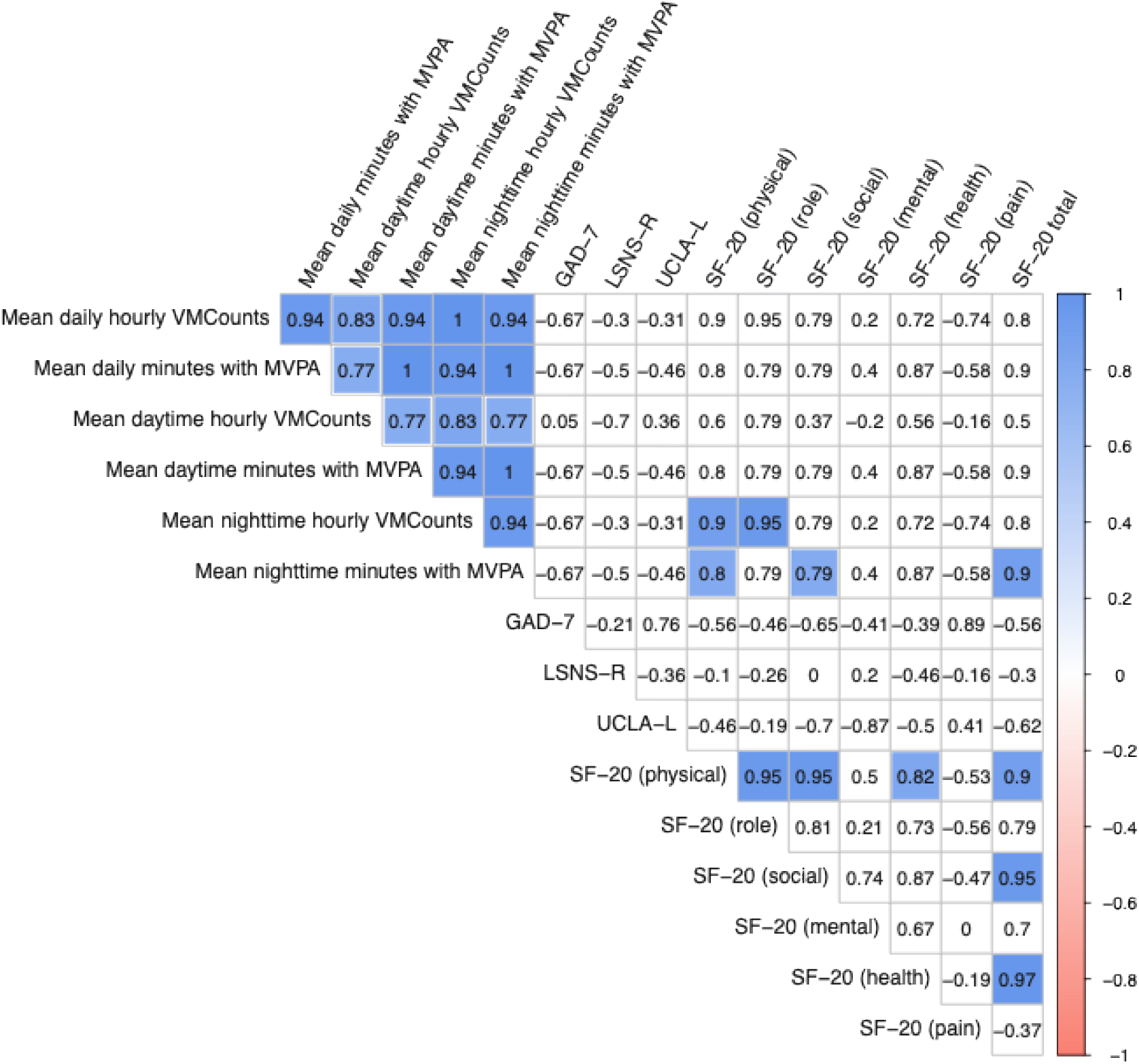
Spearman’s correlation coefficient Matrix: Relationship between mean VMCounts and mean minutes with MVPA (24-hr, daytime, nighttime) and GAD-7, LSNS-R, UCLA-L, and SF-20 subdomains in Phase A. Colored cells indicate nominal p-values <0.05.

**Supplementary Figure 6.**
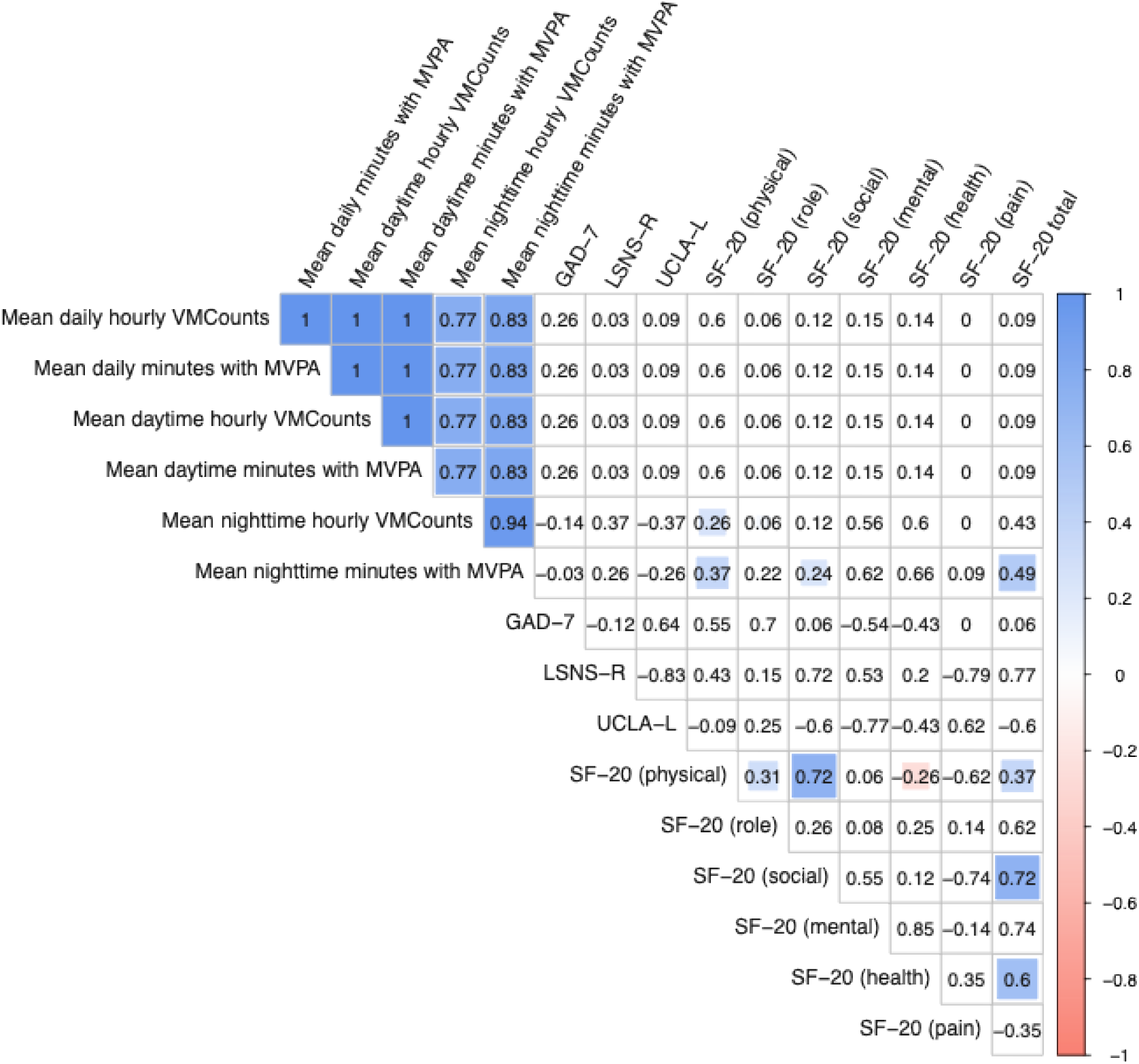
Spearman’s correlation coefficient Matrix: Relationship between mean VMCounts and mean minutes with MVPA (24-hr, daytime, nighttime) and GAD-7, LSNS-R, UCLA-L, and SF-20 subdomains in Phase B. Significant correlations are indicated by colored cells. Colored cells indicate nominal p-values <0.05.

